# An inflammatory cytokine signature helps predict COVID-19 severity and death

**DOI:** 10.1101/2020.05.28.20115758

**Authors:** Diane Marie Del Valle, Seunghee Kim-Schulze, Hsin-Hui Huang, Noam Beckmann, Sharon Nirenberg, Bo Wang, Yonit Lavin, Talia Swartz, Deepu Madduri, Aryeh Stock, Thomas U. Marron, Hui Xie, Manishkumar Patel, Oliver van Oekelen, Adeeb Rahman, Patricia Kovatch, Judith A. Aberg, Eric Schadt, Sundar Jagannath, Madhu Mazumdar, Alexander Charney, Adolfo Firpo-Betancourt, Damodara Rao Mendu, Jeffrey Jhang, David Reich, Keith Sigel, Carlos Cordon-Cardo, Marc Feldmann, Samir Parekh, Miriam Merad, Sacha Gnjatic

## Abstract

The COVID-19 pandemic caused by infection with Severe Acute Respiratory Syndrome Coronavirus 2 (SARS-CoV-2) has led to more than 100,000 deaths in the United States. Several studies have revealed that the hyper-inflammatory response induced by SARS-CoV-2 is a major cause of disease severity and death in infected patients. However, predictive biomarkers of pathogenic inflammation to help guide targetable immune pathways are critically lacking. We implemented a rapid multiplex cytokine assay to measure serum IL-6, IL-8, TNF-α, and IL-1β in hospitalized COVID-19 patients upon admission to the Mount Sinai Health System in New York. Patients (n = 1484) were followed up to 41 days (median 8 days) and clinical information, laboratory test results and patient outcomes were collected. In 244 patients, cytokine measurements were repeated over time, and effect of drugs could be assessed. Kaplan-Meier methods were used to compare survival by cytokine strata, followed by Cox regression models to evaluate the independent predictive value of baseline cytokines. We found that high serum IL-6, IL-8, and TNF-α levels at the time of hospitalization were strong and independent predictors of patient survival. Importantly, when adjusting for disease severity score, common laboratory inflammation markers, hypoxia and other vitals, demographics, and a range of comorbidities, IL-6 and TNF-α serum levels remained independent and significant predictors of disease severity and death. We propose that serum IL-6 and TNF-α levels should be considered in the management and treatment of COVID-19 patients to stratify prospective clinical trials, guide resource allocation and inform therapeutic options. We also propose that patients with high IL-6 and TNF-α levels should be assessed for combinatorial blockade of pathogenic inflammation in this disease.

## INTRODUCTION

As of late May 2020, COVID-19 disease, caused by SARS-CoV-2 infection, has resulted in more than 5,000,000 infections and 350,000 deaths worldwide. A recent study from hospitals in New York City, the epicenter of the COVID-19 pandemic in the United States, reported that of 2,634 patients hospitalized with confirmed COVID-19 between March 1 and April 4 2020, 21% of patients died^1^ which correlates with the outcomes of patients in the Mount Sinai Hospital System^2^. There are currently no curative or preventive therapies for COVID-19, highlighting the need to enhance current understanding of SARS-CoV-2 pathogenesis for the rational development of therapeutics.

Recent studies have suggested that in addition to direct viral damage, uncontrolled inflammation contributes to disease severity in COVID-19^3,4^. Consistent with this hypothesis, high levels of inflammatory markers, including CRP, ferritin, D-dimer, high neutrophil-to-lymphocyte ratio^5–7^ increased levels of inflammatory cytokines and chemokines^5,7–9^ have been observed in patients with severe diseases. Pathogenic inflammation, also referred to as cytokine storm, shares similarities with what was previously seen in patients infected with other severe coronaviruses including SARS-CoV and MERs-CoV^10^ and bears similarities with the cytokine release syndrome (CRS) observed in cancer patients treated with chimeric antigen receptor modified T cells (CAR-T)^11^. Tocilizumab, an IL-6 receptor inhibitor, is an FDA approved treatment for CRS in patients receiving CAR-T cells^12^. A number of single center studies have used IL-6 inhibitors to treat COVID-19 patients with some clinical benefits^13^, and reported failures^12^. Beyond IL-6, several cytokines have been shown to be elevated in CRS and to contribute to tissue damage. TNF-α is important in nearly all acute inflammatory reactions, acting as an amplifier of inflammation, and its blockade has been used to treat more than ten different autoimmune inflammatory diseases, suggesting that TNF-α blockade may be an interesting therapeutic approach to reduce organ damage in COVID-19 patients^14^. IL-1 is also a highly active pro-inflammatory cytokine and monotherapy blocking IL-1 activity is used to treat inflammatory diseases including rheumatoid arthritis and inherited autoinflammatory syndromes such as cryopyrin associated syndromes and has led to sustained reduction in disease severity^15^. IL-8 is a potent proinflammatory cytokine playing a key role in the recruitment and activation of neutrophils during inflammation^16^ and, given the frequent neutrophilia observed in patients infected with SARS-CoV2, it is possible that IL-8 contributes to COVID 19 pathophysiology.

To mitigate inflammation caused by SARS-CoV-2, a frenzy of immunomodulatory agents including small molecules and monoclonal antibodies targeting cytokines have rapidly been entering into clinical trials^3^ and many such FDA-approved agents are already being used routinely in the clinic in an off-label manner. Given the significant side effects associated with the use of these agents, there is an urgent need to identify biomarkers that can accurately predict which patients will decompensate due to an unchecked inflammatory response, and help guide rational targeted immunomodulatory therapeutic strategies.

In this study, we asked whether inflammatory cytokine levels can help predict disease course and outcome in COVID-19 patients. To enhance the relevance of the cytokine assays, we focused on four pathogenic cytokines, IL-6, IL-8, TNF-α, and IL-1β, with clinically available drugs to counteract them, and chose the ELLA microfluidics platform to rapidly measure them (within 3 hours), making these results potentially actionable.

We followed 1,484 patients hospitalized for suspected or confirmed COVID-19 at the Mount Sinai Health System from the day of hospitalization to the day of discharge or death. We measured serum IL-6, IL-8, TNF-α, and IL-1β levels upon admission and correlated these results with clinical and laboratory markers of disease severity and with disease outcome. We found that elevated IL-6 and TNF-α serum levels at presentation were strong predictors of disease severity and survival independently of standard clinical biomarkers of disease severity, including laboratory and clinical factors. These results suggest that multiplex cytokine profiling should be used to stratify patients and guide resource allocation and prospective interventional studies.

## RESULTS

### Cohort characteristics and cytokine ranges

We obtained laboratory and clinical information as part of standard practice from 1,484 patients with suspected or confirmed SARS-CoV-2 infection and hospitalized at the Mount Sinai Hospital System in New York City between March 21 and April 28, 2020, under expedited IRB approval. Using an emergency use approval from the New York State Department of Health, we implemented the ELLA microfluidics soluble analyte test in the clinical laboratories to measure four inflammatory cytokines known to contribute to pathogenic inflammation in CAR-T cell associated CRS – IL-6, IL-8, TNF-α and IL-1β – and assessed their correlation with severity and survival. Of the patients tested, 1,257 had a documented positive or presumptive positive SARSCoV-2 PCR test, while the remaining 167 could not be confirmed.

A total of 1953 specimens were analyzed to quantify circulating IL-6, IL-8, TNF-α and IL-1β, serum levels using the ELLA rapid detection ELISA microfluidics platform (described in Methods and in Supplemental Figure 1A-C). In most of the 1,484 patients accrued, samples were collected once, typically upon admission to the hospital (median, 1.1 day; IQR, 0.7–2.6). A subset of patients (n = 244) had repeated cytokine measurements performed more than once after admission, though for all prognostic analyses, only the first available test was used. For the entire cohort, the median time available from first cytokine test to last follow-up (i.e., date of discharge, date of death, or date still in hospital, whichever is latest) was 8 days (IQR, 2.9–14.7, up to 41 days). Patient characteristics are listed in Table 1. As reference, and to serve as control, cytokine measurements were previously performed in healthy donors and in cancer patients who either developed or did not develop CRS post-CAR-T cell therapies.

**Figure 1.**
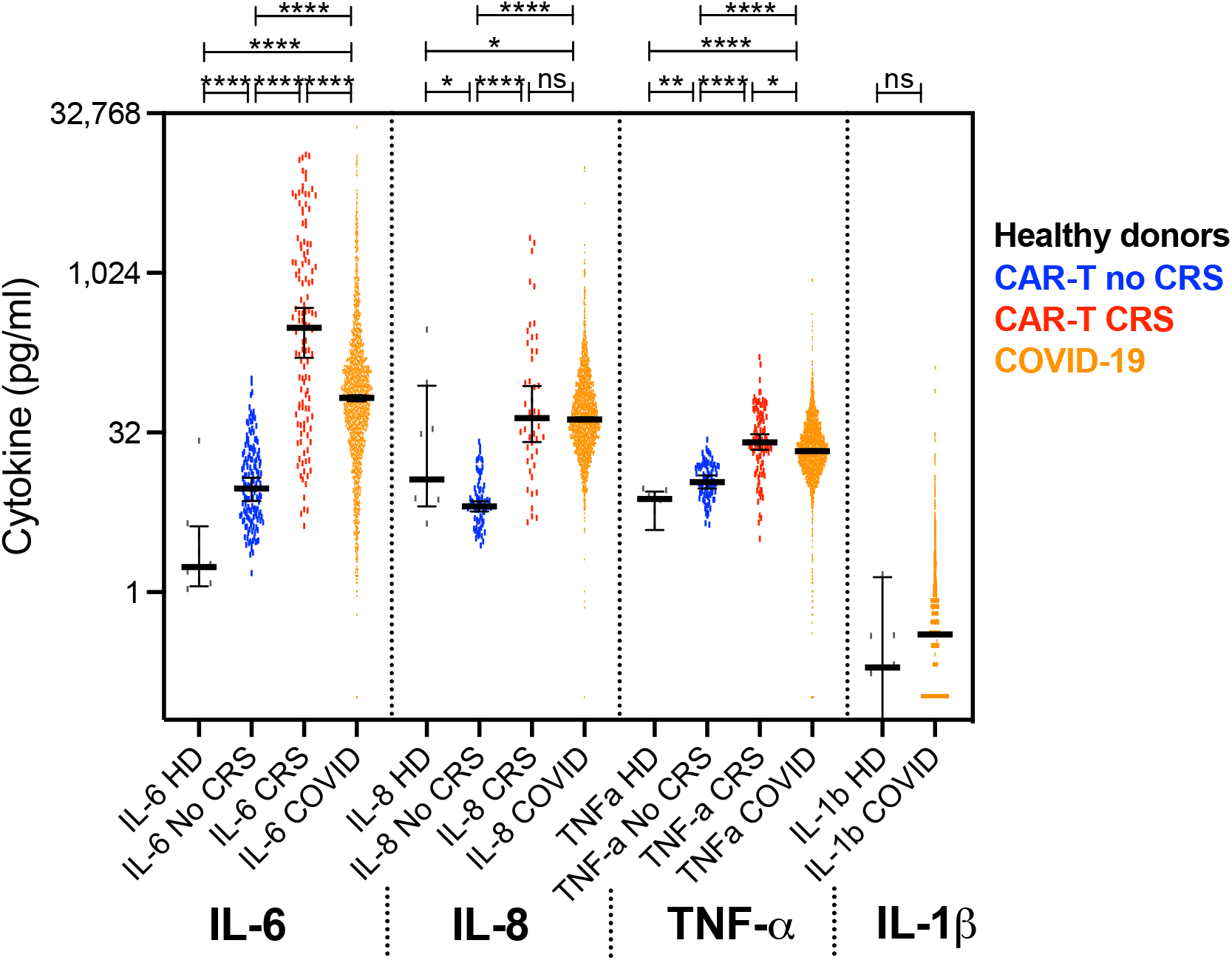
Detection range of cytokines in all tested serum samples from COVID-19 patients hospitalized at the Mount Sinai Health System (orange, n = 1959), in comparison with serum samples from healthy donors (black, n = 9) and plasma samples from multiple myeloma patients prior (blue, n = 151) and during (red, n = 121) CRS induced by CAR-T cell therapy. Heavy bars indicate median, error bars represent 95% confidence interval, each value indicated by a dot. Pairwise comparisons by Mann-Whitney t-test show significantly higher levels of IL-6, IL-8, and TNF-α in COVID-19 samples compared to samples from healthy donors of non-CRS cancer patients (****: p<0.0001, ***: p<0.001, **: p<0.01, *: p<0.05, ns: not significant). Median, mean, and range shown in Supplemental Figure 1D.

**Table 1.** Patient characteristics.

| **Table 1. Patient characteristics** | **SARS-Cov-2 PCR confirmed patients** | **SARS-Cov-2 PCR negative patients** |
| --- | --- | --- |
| Median age (IQR) – yr | 63 (53-72) | 60 (49-73) |
| Male | 787/1309 (60.1%) | 90/167 (53.9%) |
| Race/ethnicity – Hispanic | 577/1268 (45.5%) | 62/167 (37.1%) |
| Race/ethnicity – African American | 278/1268 (21.9%) | 46/167 (27.5%) |
| Race/ethnicity – White | 277/1268 (21.8%) | 43/167 (25.7%) |
| Race/ethnicity – Asian | 73/1268 (5.8%) | 5/167 (3.0%) |
| Race/ethnicity – Other | 63/1268 (5.0%) | 11/167 (6.6%) |
| Obesity (BMI≥30) | 465/1176 (39.5%) | 34/149 (22.8%) * |
| Co-morbidities – Hypertension | 420/1268 (33.1%) | 67/167 (40.1%) |
| Co-morbidities – Diabetes | 293/1268 (23.1%) | 34/167 (20.4%) |
| Co-morbidities – Chronic Kidney Disease | 167/1268 (13.2%) | 27/167 (16.2%) |
| Co-morbidities – Cancer (active) | 147/1267 (11.6%) | 37/166 (22.3%) |
| Co-morbidities – Atrial Fibrillation | 123/1267 (9.7%) | 9/167 (5.4%) * |
| Co-morbidities – Congestive Heart Failure | 69/1268 (5.4%) | 18/167 (10.8%) * |
| Co-morbidities – Asthma | 72/1268 (5.7%) | 13/167 (7.8%) |
| Co-morbidities – COPD | 44/1268 (3.5%) | 18/167 (10.8%) |
| Co-morbidities – Sleep Apnea | 50/1268 (3.9%) | 6/167 (3.6%) |
| Smoking – current | 55/965 (5.7%) | 27/143 (18.9%) * |
| Smoking – history | 293/965 (30.4%) | 53/143 (37.1%) |
| SARS-CoV-2 PCR detected | 1257/1257 (100.0%) | 0/167 (0.0%) * |
| SARS-CoV-2 Ab detected | 65/73 (89.0%) | 3/5 (60.0%) |
| Maximum Severity Score – mild/moderate | 505/1168 (43.2%) | 99/150 (66.0%) * |
| Maximum Severity Score – severe | 285/1168 (24.4%) | 22/150 (14.7%) * |
| Maximum Severity Score – severe with end organ damage | 378/1168 (32.4%) | 29/150 (19.3%) * |
| ARDS | 199/1267 (15.7%) | 11/167 (6.6%) * |
| Died within follow-up period | 269/1317 (20.4%) | 13/167 (7.8%) * |

\*Significantly different between PCR positive and negative subsets by Fisher's exact or Chi-square test. #Severity score, described in Methods, is a composite measurement for COVID-19 established by Mount Sinai infectious disease and pulmonology teams based on criteria used in the literature, and includes creatinine clearance, ALT levels, use and type of respirators/ventilators, and vasopressors. Severity score indicated here is the maximum achieved throughout the observation period, but for all other analyses, the severity score at time of first measurement was used.

We found that IL-6, IL-8, and TNF-α were significantly elevated in COVID-19 serum compared to healthy donor serum or plasma isolated from CAR-T cell treated patients with no CRS (Figure 1). The four cytokines assessed had different detection ranges, with IL-6 having the most dynamic profile, followed by IL-8, and TNF-α (Figure 1, Supplementary Figure 1D). In line with previous reports, IL-1β levels were mostly low or at the limit of detection (even though controls worked as expected, Supplementary Figure 1B). The vast majority of patients therefore presented with elevated cytokines, or cytokine storm, but in contrast to the coordinated increase in cytokines during CAR-T CRS (average Spearman r = 0.6), cytokine levels were not as highly correlated with each other in COVID-19 samples (average Spearman r = 0.4), suggesting differential patterns of cytokine expression and potentially distinct clinical presentations based on the relative profile of each independent cytokine (Supplemental Figure 1E and 1F). Since > 70% of samples analyzed for each cytokine in COVID-19 fell within CRS range based on our post-CAR-T defined cutoffs, and because we did not have an established cutoff for IL-1β, we decided to separate high vs. low values using the median for each cytokine in COVID-19 patients. The median cutoffs for further statistical analyses were > 70 pg/mL for IL-6, > 50 pg/mL for IL-8, > 35 pg/mL for TNF, and > 0.5 pg/mL for IL-1β.

### Association with demographics and comorbidities

We used the first available cytokine measurement in each patient to measure correlations with demographics and comorbidities. We hypothesized that cytokines are elevated in COVID-19 patients compared to healthy donors and non-CRS CAR-T treated patients due to SARS-CoV-2 infection. Of the 1,484 patients hospitalized with COVID-19 symptoms, 11.7% were negative for SARS-CoV-2 by PCR and therefore were excluded from further univariate analyses. It should be noted that there may have been false negative tests for SARS-CoV-2 viral detection based on subsequent tests demonstrating antibodies to SARS-CoV-2 S spike protein in 3/5 SARS-Cov-2 PCR negative patients tested. Despite similar comorbidities, cytokine levels in this subset of patients were significantly lower compared to patients that tested positive for SARS-CoV-2 infection (Figure 2A).

**Figure 2.**
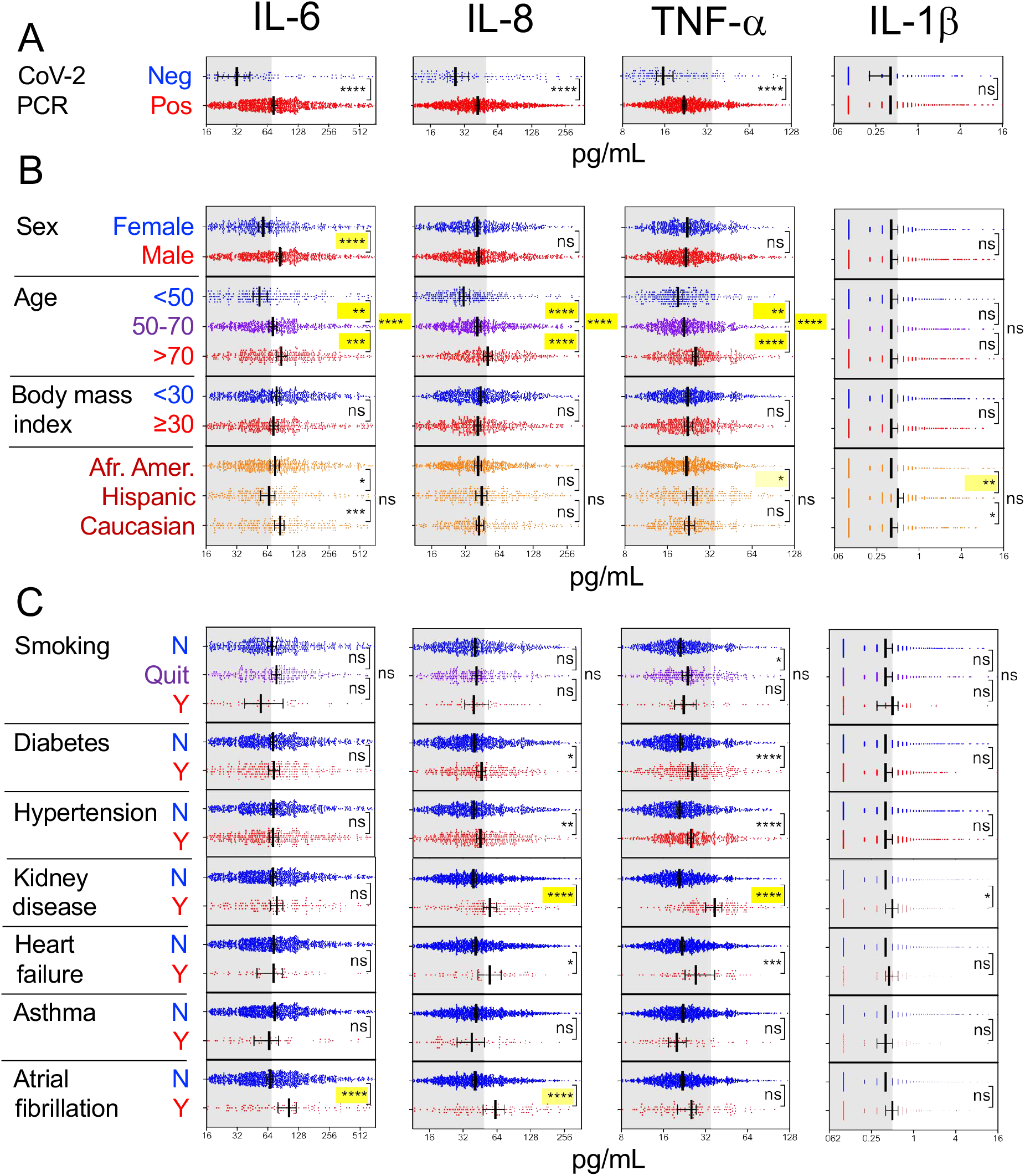
Cytokine levels observed in relation to A) SARS-CoV-2 PCR status (negative indicates patients with COVID-19-like respiratory symptoms with a negative SARS-CoV-2 PCR), B) demographics (excluding PCR negative), and C) comorbidities (excluding PCR negative, see numbers in Table 1). Scatter plots indicating individual measurements (dots), thick line at median, error bars representing 95% confidence interval, and statistical analyses by Mann-Whitney univariate t-test (****: p<0.0001, ***: p<0.001, **: p<0.01, *: p<0.05, ns: not significant). Not shown here were COPD, HIV, sleep apnea, and active cancer, which did not show any significant difference for cytokine levels. In yellow highlights are the statistical values still significant after adjustment of all demographic and comorbidity variables, with shade of yellow indicating adjusted p-value (light: *, mid: **, high: ***, saturated: ****). Grey area indicates cytokine levels below respective cutoff.

Men had higher levels of IL-6 than women, but no sex differences were observed for the other three cytokines (Figure 2B). With increased age brackets (<50, 50–70, > 70 years old), levels of IL-6, IL-8, and TNF-α increased (Figure 2B) and the same was observed for age when assessed as a continuous variable (not shown). There was no association of any cytokine measured with body mass index, however, there is an increased prevalence of obesity in our studied population. Smoking and race/ethnicity showed weak but significant univariate associations with IL-6, IL-1β, and/or TNF-α which were not confirmed after adjusting for the other covariates, except for IL-1β and TNF-α which remained significantly higher when comparing Hispanics to African Americans.

We then assessed whether cytokine levels were associated with co-morbidities listed in Table 1. We found that TNF-α and IL-8 were significantly increased in patients with chronic kidney disease (CKD), diabetes, and hypertension, while TNF-α was also increased in those with congestive heart failure, based on univariate analyses. IL-6 and IL-8 were elevated in patients with a history of atrial fibrillation. No associations were found between cytokines and active cancer, asthma, COPD, HIV, and sleep apnea (not shown).

Using multivariable regression models, we confirmed that CKD was the only comorbidity significantly associated with elevated cytokine levels, while elevated TNF-α in patients with diabetes and hypertension were explained by other variables. From demographics, age and sex (for IL-6) remained significantly associated with cytokine levels as seen in univariate analyses. Therefore, we included demographics and comorbidities as confounding variables in subsequent analyses. Time to ELLA testing from time of hospital admission did not impact the cytokine levels, meaning whether patients were tested for cytokines immediately upon admission or later (as will be discussed in the longitudinal follow-up section) did not impact the level of cytokines. Therefore, this time difference was not considered as a potential confounder.

### Association between cytokines and risk of death

Next, we considered factors affecting survival defined as time to death and censored regardless of cytokines in the overall cohort with univariate Kaplan-Meier analyses. We found that only age and CKD were significantly associated with increased risk of death from COVID-19. We evaluated whether cytokines could distinguish patients based on overall survival and disease severity after COVID-19 hospitalization. Stratifying patients by cytokine levels of high vs. low using the cutoffs described in the statistical analysis section, we found that each cytokine could predict the overall survival of patients, based on the first available measurement following hospital admission. Each cytokine was independently predictive of overall survival, after adjusting for demographics and comorbidities, i.e., sex, age, race/ethnicity, smoking, CKD, hypertension, asthma, congestive heart failure (Figure 3).

**Figure 3.**
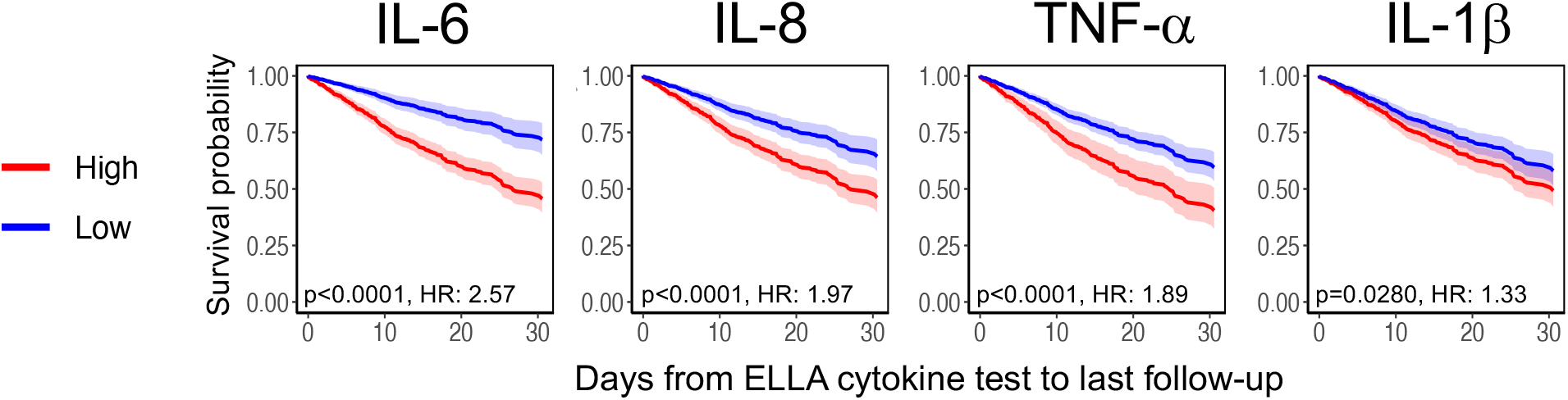
Survival curves based on each cytokine measured, after multiple variable adjustments for sex, age, race/ethnicity, smoking, CKD, hypertension, asthma, and congestive heart failure (n = 1246). Cox regression model showing overall survival with confidence intervals for each cytokine based on time from ELLA cytokine test to last follow-up date (discharge, death, or still in hospital, whichever comes last), with significance indicated by p value and hazard ratio (HR). There was worse survival if cytokines are high (red, above cutoffs of 70 pg/mL for IL-6, 50 pg/mL for IL-8, 35 pg/mL for TNF-α, and 0.5 pg/mL for IL-1β) vs. low (blue, below cutoffs).

When considering all cytokines together in the model, for their relative independence, all but IL-1β remained significant, even after adjustment for demographics and comorbidities. This confirmed the relative independence of each cytokine tested, with only age (50–70 vs. > 70, HR 0.55 [0.42–0.72]; <50 vs. > 70: 0.24 [0.15–0.41]), IL-6 (HR 2.27 [1.65–3.12], IL-8 (HR 1.48 [1.11-1.97]), and TNF-α (HR 1.50 [1.10–2.05]) remaining significantly associated with decreased survival after adjustments (respectively p<0.0001, p<0.0001, p = 0.0076, p = 0.0105). As additional validation, we also performed this analysis using a competing risk model, in which patients discharged alive were considered competing events and patients in hospital were censored, and found the same conclusion, where high IL-6, IL-8, and TNF-α remained significantly associated with worse outcome regardless of demographics and comorbidities (not shown). We used the competing risk model in the next analysis,

### Using cytokines to complement risk stratification

Next, we asked whether cytokines were of value for risk stratification and survival, independently of known laboratory and clinical severity metrics (i.e., temperature, O_2_ saturation, respiratory rate, severity score as defined in Methods). We first tested whether the 4 tested cytokine levels were associated with known inflammation markers CRP, D-dimer, and ferritin, and found strong correlations in all cytokine with each measurement, with IL-6 and IL-1β additionally associated with fever (Figure 4A). In addition, IL-6 and IL-8 levels were closely correlated with severity scale, which takes into account lung imaging and use of ventilation and end organ damage, while TNF-α did not distinguish moderate vs. severe COVID-19 presentation, but instead was only increased with end organ damage. Looking at the predictive value of cytokines on survival after adjusting for levels of CRP, D-dimer, ferritin, and all comorbidities, IL-6 and IL-8 remained independently predictive of survival, therefore showing additive value to these known markers (not shown). However, when including additional severity metrics, severity scoring, IL-8 was no longer independent from other variables, likely because it was eclipsed by stronger measurements which follows a similar pattern (Supplementary Table S1). We then investigated correlations of cytokines with an additional series of well-established markers of inflammation, renal function, myocardial strain, and respiratory distress for their impact on survival within this cohort. Using unsupervised analyses, neutrophils, white blood cells, CRP, ferritin, D-dimer, LDH, and low O2 saturation were co-clustering with all cytokines except TNF-α, which was more closely correlated with markers of tissue damage like creatinine (Supplementary Figure 2). Selecting the most informative variables using a backward elimination process to define each of the available measurements to be used as confounding factors in a competing risk regression analysis of survival along with the cytokines, we found severity score, O_2_ saturation, platelets, low albumin, systolic blood pressure, D-dimer, albumin, calcium, chloride, and platelet count remaining. Remarkably, even when using these measurements as variables to adjust when assessing the predictive value of cytokines on survival in the competing risk regression analysis, we found that IL-6 and TNF-α remained significantly associated with a worse prognosis (Figure 4B). Therefore, we conclude that IL-6 and TNF-α are independently predictive of patient outcomes in terms of both disease severity and survival (Supplementary Table S2). Even after stratifying for risk factors with the strongest p value, i.e., severity score, O_2_ saturation, and age, IL-6 and TNF-α remained independently predictive of survival (Figure 4C and Supplementary Table S3).

**Figure 4.**
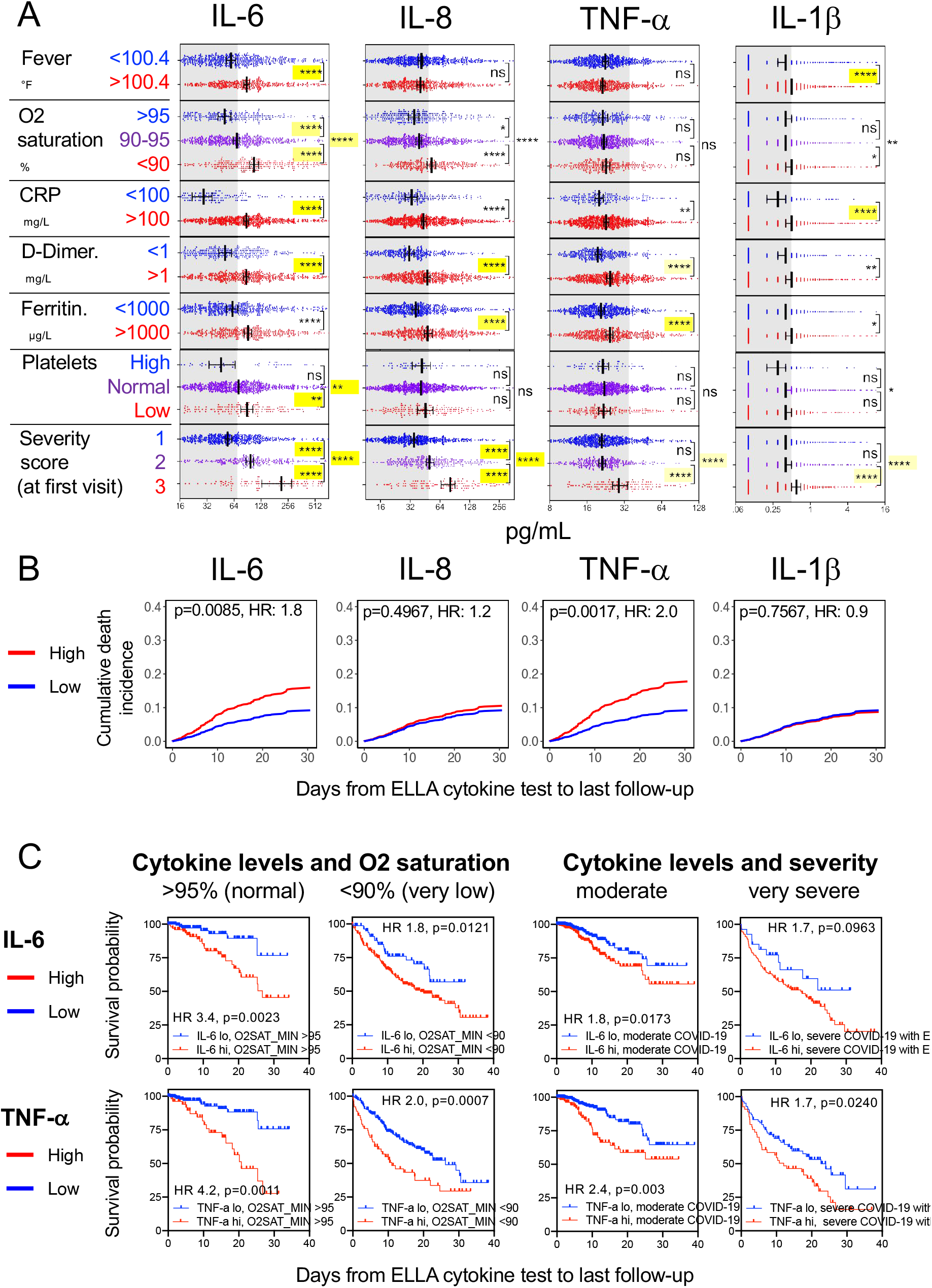
Correlation of cytokine levels with established inflammatory and severity measurements. A. Correlation of each cytokine with each metric, using same univariate and multivariate analyses as in figure 2 legend. B. Competing risk analysis (n = 671) showing survival differences by IL-6 and TNF-α levels, after adjusting the following variables: IL-6, IL-8, TNF-α, IL-1β, age, sex, race/ethnicity, smoking status, asthma, atrial fibrillation, cancer, CHF, CKD, COPD, diabetes, HPN, sleep apnea, severity, systolic blood pressure max, O2 saturation min, D-dimer, albumin, calcium, chloride, and platelet count. C. Kaplan-Meier univariate analyses of survival by IL-6 and TNF-α levels in patients with normal (n = 257) vs. very low (n = 287) O2 saturation, or in patients with moderate (n = 588) vs. severe CODID-19 with end organ damage (n = 136), as measured at the first available test.

**Supplementary Table S1.** Competing risk model with four cytokines, demographics, comorbidities, and laboratory measurements. After adjustments, IL-6 and TNF-α remain significantly predictive.

SUPPLEMENTARY TABLES AND FIGURES
| Competing risk model with multiple variable adjustments |  |  |  |
| --- | --- | --- | --- |
| Variables | Levels | HR (95% CI) | Overall P-value |
| TNF-α | High | 1.85 (1.21, 2.82) | **0.0046** |
|  | Low | Ref |  |
| IL-6 | High | 2.06 (1.33, 3.18) | **0.0011** |
|  | Low | Ref |  |
| IL-8 | High | 1.37 (0.94, 2.00) | 0.0973 |
|  | Low | Ref |  |
| IL-1β | High | 0.95 (0.68, 1.34) | 0.7671 |
|  | Low | Ref |  |
| CRP | High | 1.27 (0.85, 1.90) | 0.2366 |
|  | Low | Ref |  |
| D-DIMER | High | 0.88 (0.55, 1.42) | 0.6040 |
|  | Low | Ref |  |
| FERRITIN | High | 0.86 (0.59, 1.24) | 0.4128 |
|  | Low | Ref |  |
| SEX | Female | 1.23 (0.87, 1.75) | 0.2494 |
|  | Male | Ref |  |
| AGE | 50-70 | 0.57 (0.40, 0.82) | **0.0003** |
|  | <50 | 0.31 (0.16, 0.58) |  |
|  | >70 | Ref |  |
| RACE/ETHNICITY | NH African American | 0.73 (0.42, 1.28) | 0.6318 |
|  | Hispanic | 0.99 (0.64, 1.54) |  |
|  | Other | 0.92 (0.51, 1.67) |  |
|  | NH White | Ref |  |
| BMI | BMI>30 | 0.99 (0.67, 1.46) | 0.9643 |
|  | BMI≤30 | Ref |  |
| DIABETES | Yes | 0.80 (0.54, 1.20) | 0.2824 |
|  | No | Ref |  |
| HYPERTENSION | Yes | 1.08 (0.71, 1.65) | 0.7222 |
|  | No | Ref |  |
| SMOKING STATUS | Active smoker | 1.40 (0.63, 3.10) | 0.4204 |
|  | Quit | 1.43 (0.91, 2.23) |  |
|  | Unknown | 1.06 (0.67, 1.67) |  |
|  | Non-smoker | Ref |  |
| CKD | Yes | 1.84 (1.07, 3.18) | **0.0274** |
|  | No | Ref |  |
| ASTHMA | Yes | 1.03 (0.43, 2.47) | 0.9523 |
|  | No | Ref |  |
| CHF | Yes | 0.72 (0.26, 2.03) | 0.5365 |
|  | No | Ref |  |
| COPD | Yes | 0.84 (0.33, 2.16) | 0.7160 |
|  | No | Ref |  |
| SLEEP APNEA | Yes | 1.59 (0.72, 3.51) | 0.2499 |
|  | No | Ref |  |
| ATRIAL FIBRILLATION | Yes | 0.91 (0.53, 1.56) | 0.7184 |
|  | No | Ref |  |
| CANCER | Yes | 1.73 (1.05, 2.84) | **0.0315** |
|  | No | Ref |  |
| SEVERITY SCORES | Severe | 3.20 (2.05, 4.99) | **<.0001** |
|  | Severe with EOD | 5.69 (3.38, 9.60) |  |
|  | Moderate | Ref |  |

**Supplementary Table S2.**
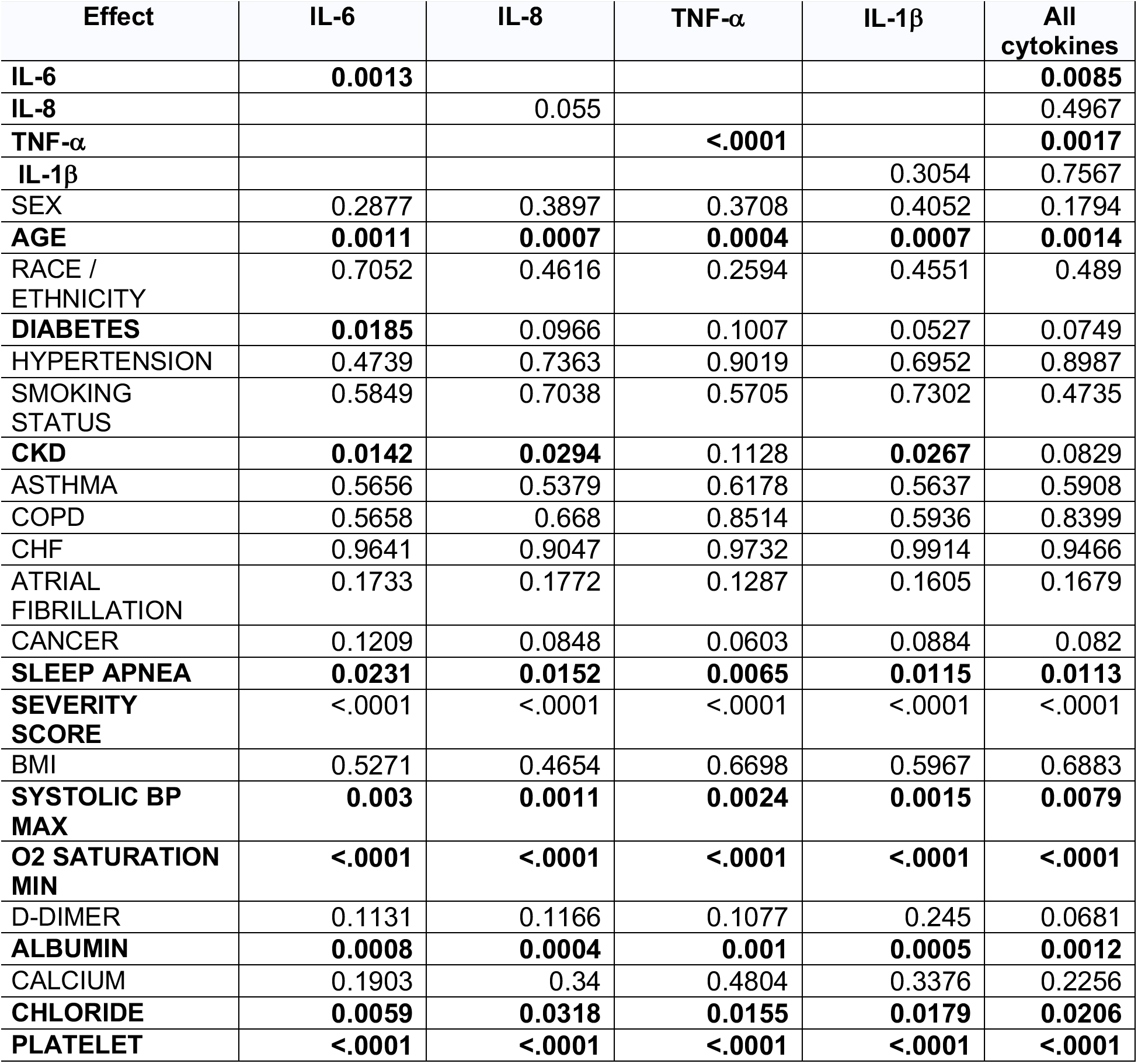
Type 3 test of competing risk survival analysis for cytokine tested in ELLA and adjusted for comorbidities and laboratory/clinical metrics. After adjustment, IL-6 and TNF-a remain significantly predictive of survival based on a competing risk model.

| Competing risk models adjusting for multiple variables (Overall P-value) |  |  |  |  |  |
| --- | --- | --- | --- | --- | --- |
| Effect | IL-6 | IL-8 | TNF-α | IL-1β | All cytokines |
| IL-6 | **0.0013** |  |  |  | **0.0085** |
| IL-8 |  | 0.055 |  |  | 0.4967 |
| TNF-α | | | **<.0001** | | **0.0017** |
| IL-1β | | | | 0.3054 | 0.7567 |
| SEX | 0.2877 | 0.3897 | 0.3708 | 0.4052 | 0.1794 |
| AGE | **0.0011** | **0.0007** | **0.0004** | **0.0007** | **0.0014** |
| RACE / ETHNICITY | 0.7052 | 0.4616 | 0.2594 | 0.4551 | 0.489 |
| DIABETES | **0.0185** | 0.0966 | 0.1007 | 0.0527 | 0.0749 |
| HYPERTENSION | 0.4739 | 0.7363 | 0.9019 | 0.6952 | 0.8987 |
| SMOKING STATUS | 0.5849 | 0.7038 | 0.5705 | 0.7302 | 0.4735 |
| CKD | **0.0142** | **0.0294** | 0.1128 | **0.0267** | 0.0829 |
| ASTHMA | 0.5656 | 0.5379 | 0.6178 | 0.5637 | 0.5908 |
| COPD | 0.5658 | 0.668 | 0.8514 | 0.5936 | 0.8399 |
| CHF | 0.9641 | 0.9047 | 0.9732 | 0.9914 | 0.9466 |
| ATRIAL FIBRILLATION | 0.1733 | 0.1772 | 0.1287 | 0.1605 | 0.1679 |
| CANCER | 0.1209 | 0.0848 | 0.0603 | 0.0884 | 0.082 |
| SLEEP APNEA | **0.0231** | **0.0152** | **0.0065** | **0.0115** | **0.0113** |
| SEVERITY SCORE | <.0001 | <.0001 | <.0001 | <.0001 | <.0001 |
| BMI | 0.5271 | 0.4654 | 0.6698 | 0.5967 | 0.6883 |
| SYSTOLIC BP MAX | **0.003** | **0.0011** | **0.0024** | **0.0015** | **0.0079** |
| O2 SATURATION MIN | **<.0001** | **<.0001** | **<.0001** | **<.0001** | **<.0001** |
| D-DIMER | 0.1131 | 0.1166 | 0.1077 | 0.245 | 0.0681 |
| ALBUMIN | **0.0008** | **0.0004** | **0.001** | **0.0005** | **0.0012** |
| CALCIUM | 0.1903 | 0.34 | 0.4804 | 0.3376 | 0.2256 |
| CHLORIDE | **0.0059** | **0.0318** | **0.0155** | **0.0179** | **0.0206** |
| PLATELET | **<.0001** | **<.0001** | **<.0001** | **<.0001** | **<.0001** |

**Supplementary Table S3.** After stratification by the most predictive variables (O2 saturation, age group, and severity score), the independent predictive value of IL-6 and TNF-α remains, and IL-8 returns to significance. The estimates were obtained using SAS PHREG procedure and STRATA statement.

| Stratification | Effect | HR (95% CI) | P-value |
| --- | --- | --- | --- |
| O2 saturation | **TNF-α** | 2.12 (1.45, 3.10) | **0.0001** |
|  | **IL-6** | 1.91 (1.24, 3.93) | **0.0031** |
|  | **IL-8** | **1.49 (1.03, 2.17)** | **0.0353** |
| | IL-1β | 1.22 (0.86, 1.73) | 0.2639 |
| Age Group | **TNF-α** | 1.93 (1.34, 2.79) | **0.0005** |
|  | **IL-6** | 1.96 (1.30, 2.97) | **0.0015** |
|  | **IL-8** | **1.46 (1.01, 2.11)** | **0.0472** |
| | IL-1β | 1.20 (0.85, 1.68) | 0.3057 |
| Severity Score | **TNF-α** | **1.99 (1.38, 2.88)** | **0.0002** |
|  | **IL-6** | **1.96 (1.30, 2.98)** | **0.0015** |
|  | **IL-8** | **1.46 (1.01, 2.12)** | **0.0455** |
| | IL-1β | 1.19 (0.85, 1.67) | 0.3182 |

\*Each model includes one cytokine
\*Covariates: sex, age, race/ethnicity, diabetes, hypertension, smoking status, CKD, asthma, congestive heart failure (CHF), atrial fibrillation, chronic obstructive pulmonary disease (COPD), cancer, severity score, body mass index (BMI), systolic blood pressure max, O2 saturation min, D-dimer, albumin, calcium, chloride, platelets.
\*O2 saturation min is not in the covariates of the model stratified by O2 saturation
\*Age is not in the covariates of the model stratified by age group
\*Severity score is not in the covariates of the model stratified by Severity Score

### Effect of medication and treatment on cytokine levels

Although our data does not demonstrate a causative role for IL-6 and TNF-α on disease outcome, we wanted to shed light on the effect of various treatments on measured cytokines, as potential mitigation strategies should there be a pathogenic effect from these inflammatory agents. From a subset of 244 patients with more than one ELLA cytokine assay performed, and by mapping time from treatment start to first ELLA test, we were able to assess the effect of various treatments and experimental drugs on cytokine levels (Figure 5). Interestingly, off-label treatment with the anti-IL-6R monoclonal antibody tocilizumab, which was given in a subset of patients with progressive respiratory failure with marked systemic inflammation, showed that patients who received this drug started with elevated IL-6 levels, and then had a transient increase in serum IL-6, which has previously been explained by disrupted clearance following drug saturation of the IL-6 receptor^17^. This transient elevation was only observed for IL-6, not IL-8, while TNF-α appeared gradually decrease following therapy. Other treatments such as corticosteroids and remdesivir showed, respectively, a rapid and gradual reduction in IL-6 over time compared to patients who did not receive these drugs, but no effect on TNF-α, while hydroxychloroquine, acetaminophen, or anti-coagulants did not clearly appear to alter cytokine levels. The clinical outcome of patients stratified based on IL-6 levels will need to be confirmed in prospective randomized trials.

**Figure 5.**
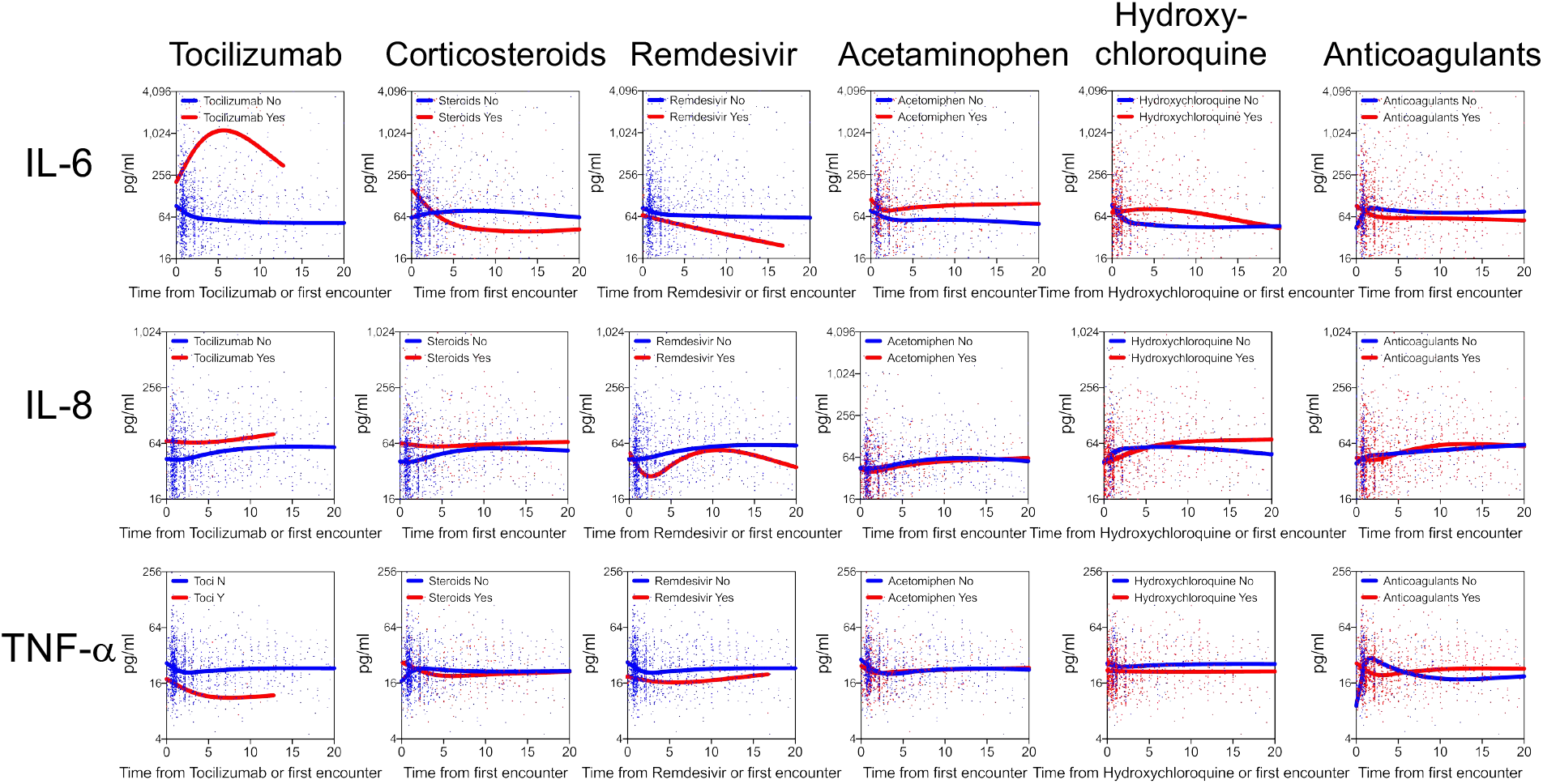
Effect of treatments on IL-6 (top row) and IL-8 (middle row) and TNF-α (bottom row). Lines indicate the best fit curve by smoothed spline of the longitudinal and unique time point distribution of each cytokine level based on time from either first encounter or treatment start. Out of 1,670 samples representing various time points of 1,315 patients with available information, the number of those from patients who received tocilizumab, corticosteroids (any of prednisone, methylprednisolone, dexamethasone), remdesivir, acetaminophen, hydroxychloroquine, and/or anticoagulants (apixaban, enoxaparin, heparin, rivaroxaban) was 73, 305, 76, 620, 1333, and 1113, respectively.

## DISCUSSION

As COVID-19 started overwhelming the health systems around the world, we aimed to understand the role of inflammatory cytokines on disease course and outcome. We established a rapid multiplex cytokine test to measure IL-6, TNF-α, and IL-1β, as known markers of inflammation and organ damage, along with CXCL8/IL-8 because of its potent role in the recruitment and activation of neutrophils, commonly elevated in COVID-19 patients. Importantly, drugs blocking these cytokines are either FDA approved or in clinical trials. Studying over 1,400 hospitalized patients in a month, we established that COVID-19 is associated with high levels of all four cytokines at presentation. Importantly, our observations indicate that cytokine patterns are predictive of COVID-19 survival and mortality, independently of demographics, comorbidities, but also of standard clinical biomarkers of diseases severity, including laboratory and clinical factors. We found that IL-6 was one of the most robust prognostic markers of survival, eclipsing or outperforming CRP, D-dimer, and ferritin after adjusting for the demographic features and comorbidities. It remained independently associated with severity and predictive of outcome when including information about ventilation and end organ damage. Furthermore, elevated TNF-α, known to contribute to organ damage, was also a strong predictor of poor outcome even after adjusting for other risk factors such as age, sex, hypoxia, disease severity scoring based on clinical assessment, and IL-6. Our cytokine panel also included IL-8, which showed association with survival time even though it was eclipsed by other severity factors after multivariate adjustments, and IL-1β which was poorly detected and as a result had only marginal predictive value. Although classic markers used routinely to determine inflammation and severity were still useful to stratify patients on their own, when combined in multivariate analyses, many were no longer significant likely due to collinearity, while IL-6 and TNF-α remained independently predictive of outcome. Both overall survival and for competing risk models used here consistently showed the significant prognostic value of TNF-α and IL-6 when all tested cytokines were in the model, along with demographics, comorbidities, and other clinical and laboratory measurements, highlighting the robustness of our findings. Interestingly, the COVID-19-related cytokine response was quite distinct from the traditional cytokine storm associated with sepsis and CAR-T cells, with sustained elevated cytokine levels over days and weeks, and relative absence of coordination among them. This raises possibilities of honing on mitigation strategies with anti-cytokine treatments, though which one(s) and the window of opportunity for their use remain to be established. Guiding such therapies based on mechanistic association with cytokine levels could provide a rational approach.

Trials to block IL-6 signaling with already FDA-approved drugs have been launched across the world and some clinical benefits have been seen in a subset of patients in small single center observational studies^13,18^. In contrast, interim analysis of randomized trials with sarilumab versus placebo identified potential benefit only in patients with severe but not moderate disease^19^. There are no available data correlating levels of IL-6 and response to treatment, and none of the current studies have used cytokine profiling as part of their inclusion criteria. It is possible that patients with moderate disease and high IL-6 levels will best benefit from cytokine blockade. There is also a need to evaluate the effect of anti-TNF-α therapy on its own in COVID-19. Since IL-6 and TNF-α appear to be independent variables, studies with combination regimen blocking both cytokines would be important to consider for added clinical efficacy.

Early cytokine measurements are reliable predictors of outcome and therefore raise the critical importance of using serum cytokine levels for treatment decisions. The predictive value of these cytokines may help to inform therapeutic interventions to determine which individuals are likely to develop respiratory failure, end organ damage, and death. Cytokine levels can serve to evaluate appropriateness for pharmacotherapy with available or experimental agents and to select optimal trial designs to disrupt the underlying inflammatory milieu. A prediction model built on cytokine levels early in disease may serve to inform health care allocation and prioritization of individuals at highest risk.

Our current efforts are to build such a predictive model from our current study using a validation cohort of more than 700 patients admitted from April 20, 2020 onward, and to supplement it with data from higher dimensional assays such as Olink proximity extension assay and Somalogics aptamer platform^20^. which can assay hundreds to thousands of soluble analytes from serum or plasma. The most informative dimensions from these assays could then be carried back into the rapid 4–8 plex ELLA cytokine detection system for clinical decision making, in addition to IL-6 and TNF-α. We believe that these practices will bring cytokine measurements to standard of care in prognosticating and monitoring of patients with COVID-19.

## METHODS

### ELLA cytokine test

The ELLA platform is a rapid cytokine detection system based on four parallel singleplex microfluidics ELISA assays run in triplicate within cartridges following manufacturer’s instructions. We first validated IL-6, IL-8, and TNF-α detection by ELLA at the Mount Sinai Human Immune Monitoring Center using plasma from multiple myeloma patients undergoing immunotherapies such as CAR-T cells and bispecific antibodies, known to elicit cytokine release storm. Analytical validation (Supplementary Figure 1A-C) was performed using both reference cytokine controls and biological replicates across different lots of cartridges, the reproducibility was > 95%, with an intra-assay CV of 0.8%, and an inter-assay CV of 0.4–0.8% for analytes in the high detection range (>250 pg/mL), and a CV of 2.6–4.2% for analytes in the lowest detection range (5–50 pg/mL). Serum and plasma appear to be equivalent for detection of these cytokines. In March 2020, as the number of COVID-19 cases was increasing in New York City, we transferred the ELLA methodology to the Mount Sinai Hospital Center for Clinical Laboratories, which allowed the ELLA cytokine test to be coded into our electronic health record ordering system as part of a COVID-19 diagnostic panel.

### Patient information and data source

This study was reviewed and approved by the Mount Sinai Institutional Review Board. A waiver of informed consent was obtained to query the patient electronic medical record. Samples for the RT-PCR SARS-CoV-2 lab test were collected via nasopharyngeal or oropharyngeal swab at one of 53 different Mount Sinai locations, representing outpatient, urgent care, emergency and inpatient facilities. Blood specimen for ELLA were collected via venipuncture within the MSHS. All specimen and imaging were collected as part of standard of care.

Between March 21 and April 28, 1,484 patients hospitalized with suspicion of COVID-19 were tested for SARS-CoV-2 viral infection status by PCR and for the ELLA cytokine panel, and we obtained routine laboratory measurements, blood counts, and as part of their medical care. Patients were identified by querying the pathology department electronic database for individuals with both SARS-CoV-2 PCR-based testing and ELLA cytokine panel. Cytokine data was obtained from pathology department electronic databases and clinical and demographic data was supplemented with information from the Mount Sinai Data Warehouse, obtained from EPIC electronic medical records (Verona, WI). Clinical follow-up data was collected up to May 7, 2020. Two investigators (D.M.D.V. and S.G.) independently compiled all clinical and laboratory information from these various sources, and compared them with near total matches. Differences were adjudicated based on individual patient chart review, and were explained by either missing or updated information from the data warehouse.

### Variables

Our dataset included 3 broad classes of variables: (1) demographic variables (age, sex, race, ethnicity and smoking status); (2) clinical variables for each day of hospital encounter (body mass index (BMI), heart rate, temperature, respiratory rate, oxygen (O_2_) saturation, systolic blood pressure, diastolic blood pressure, admission status, discharge status, and deaths; and (3) comorbid conditions (chronic kidney disease (CKD), asthma, chronic obstructive pulmonary disease (COPD), hypertension, obesity, diabetes, human immunodeficiency virus (HIV), sleep apnea, and cancer). All 3 categories were obtained from the patients’ electronic medical record with comorbid conditions defined as an active ICD code and vital signs recorded for each patient’s given encounter.

### Determining COVID-19 Disease Severity

A severity scale for COVID-19 was devised by pulmonologists at Mount Sinai based on literature^21^ and clinical practice, which defined categories as follows: 1) mild/moderate COVID-19, based on normal/abnormal (<94%) oxygen saturation, respectively, or pneumonia on imaging; 2) severe COVID-19, based on use of high-flow nasal cannula (HFNC), non-rebreather mask (NRB), bilevel positive airway pressure (BIPAP) or mechanical ventilation, and no vasopressor use, and based on creatinine clearance (CrCl) > 30 and ALT < 5x upper limit of normal; and 3) Severe COVID-19 with end organ damage, based on use of HFNC, NRB, BIPAP or mechanical ventilation with use of vasopressors, or based on CrCl <30, new renal replacement therapy (hemodialysis/continuous veno-venous hemofiltration) or ALT > 5x ULN. Clinical notes and imaging reports were reviewed in effort to establish the patient’s COVID-19 disease severity over time. Using a bag-of-words approach to vectorize both clinical notes and image reports, vectors were derived from the chest x-ray imaging report, in order to reflect the presence of viral pneumonia and worsening respiratory symptoms. Intubation status and oxygen therapy modality were obtaining by examining the patient clinical notes. The use of endotracheal tube, bilevel positive airway pressure, continuous positive air pressure, high-flow nasal cannula, mechanical ventilator, and/or supplemental oxygen greater than FiO_2_ 70% was associated with severe COVID-19. End organ damage was defined by an alanine aminotransferase level of greater than five times the upper limit of normal, creatinine clearance less than 30, use of vasopressors, and/or new renal replacement therapy.

### Sample size and power consideration

The primary endpoint is the hazard ratio of death between the high and low levels of the cytokines (TNF-α, IL-6, IL-8, and IL-1β). We tested the hypothesis for H_0_: HR_TNF-α_ = HR_IL-6_ = HR_IL-8_ = HR_IL-1β_ = 1 against H_a_: any HR≠1. In this study, the event of interest is death. A two-sided log-rank test with an overall sample size of 674 patients (337 in each group) achieves 80% power at a 0.05 type I error to detect a hazard ratio of 2.29 when the proportion of death in the low group is 20%. The study lasted for 40 days with an accrual time of 30 days and a median follow-up time of 9 days. The proportion of patients who are still in the hospital during the study period is 10%. The sample size calculation was conducted using PASS software. The primary analyses assessed the impact of the cytokine levels after adjusting for patients’ demographics and comorbidities. The patients eligible for further analyses were those with cytokine tests on the date or before the date of last follow-up (n = 1,298), which ensured the power ≥ 80% to test the primary hypothesis after adjusting for the covariates. Approximately 5% of patients had missing values in baseline demographics and comorbidities, and the cohort with complete cases was used for the primary analyses. The secondary analyses assessed the impact of the cytokines on the event of death after further adjusting for the clinical severity metrics. The analysis procedures were described in the statistical analysis section.

### Statistical analysis

Patients’ characteristics were summarized using the standard descriptive statistics: median /interquartile range (IQR) for continuous variable, and count/percent for categorical variables. Distributions of the cytokine values were assessed and log-2 transformed to render the parametric statistical analyses. The cytokines were then categorized at the level of 70 pg/mL, 50 pg/mL, 35 pg/mL, 0.5 pg/mL, 100 mg/L, 1,000 µg/L, and 1 mg/L for IL-6, IL-8, TNF-α, IL-1β, CRP, ferritin, and D-dimer, respectively. These stringent cutoffs for cytokines were decided based on the median distribution within COVID-19 samples, while those for elevated inflammatory markers were based on 2–3x the upper limit of normal detection. The univariate analyses assessed the association of the cytokines and laboratory tests with patients’ characteristics using Mann-Whitney U test, Kruskal Wallis test and Spearman’s rank correlation test as appropriate. Additionally, Deming regressions and Spearman correlation coefficients were calculated for correlations between the cytokines and laboratory tests. We used multivariable linear regression models to test the association of the cytokine values with patients’ demographics and comorbidities^22^. The Kaplan-Meier plots along with the log-rank test were conducted to assess the differences in survival probabilities between the high and low levels of each cytokine across the follow-up time frame, which was calculated from the date of cytokine testing to date of death, discharge or end of follow-up period as appropriate^23^. The Cox Proportional Hazard Model was used to estimate the hazard of death adjusting for the covariates (e.g., patients’ demographics, comorbidities, laboratory test results), which were determined by the backward elimination method^24,25^. We assessed the survival model, censoring patients discharged alive and in hospitals. The competing risk model, in which death was the event of interest, the live discharge was the competing event, and inpatients were censored^26,27^, was also fitted as a sensitivity analysis. The predicted survival probabilities and cumulative incidence curves were provided. The analyses were performed using GraphPad Prism 8.4.2., SAS 9.4, and R v.3.6.3.

## Data Availability

The datasets analyzed during the current study are not publicly available due to United States Federal Health Insurance Portability and Accountability Act (HIPAA) compliance. A de-identified dataset may be available from the corresponding author on reasonable request.

## ACKNOWLEDGEMENTS

Authors thank Nicolas Fernandez for help with the clustergrammer tool, Kevin Tuballes and other members of the Human Immune Monitoring Center for help with specimen handling. Authors wish to acknowledge Rajiv Pande and Martin Putnam at Bio-techne for helping to provide instruments and assay kits for ELLA testing in a CLIA environment in the timeliest possible way during the health crisis; and Rob Hyland from Gilead for allowing to permission to use the remdesivir-related data.

S.G., D.M.D.V.. S.K.-S., P.K., A.R. and M.M. were supported by NCI U24 grant CA224319. S.G. is additionally supported by grants U01 DK124165 and P01 CA190174. M.M. was supported by the fast-grant fund. The Human Immune Monitoring Center and the Institute for Healthcare Delivery Science received support from Cancer Center P30 grant CA196521.

S.G. reports consultancy and/or advisory roles for Merck, Neon Therapeutics and OncoMed and research funding from Bristol-Myers Squibb, Genentech, Immune Design, Agenus, Janssen R&D, Pfizer, Takeda, and Regeneron.

## AUTHOR CONTRIBUTIONS

DMDV and SKS contributed equally.

HHH and NB contributed equally.

DMDV, SKS, AR, JAA, AFP, DRM, JJ, SJ, AFP, DRM, JJ, DR, CCC, SP, MiM, and SG contributed to study concept and design.

DMDV, SKS, HHH, NB, SN, BW, YL, TS, DM, AS, TUM, OVO, AR, PK, JAA, ES, SJ, MaM, AC, AFP, DRM, JJ, DR, KS, MF, CCC, SP, MiM, and SG contributed to literature search, writing the manuscript, and data interpretation.

DMDV, SKS, HHH, NB, SN, BW, YL, TS, DM, AS, TUM, XH, MP, OVO, AR, PK, MaM, AC, AFP, DRM, JJ, DR, KS, CCC, SP, MiM, and SG participated in data collection and data analysis.

DMDV, SKS, HHH, NB, MaM, KS, and SG made figures and tables.

---

**Supplementary Figure S1.**
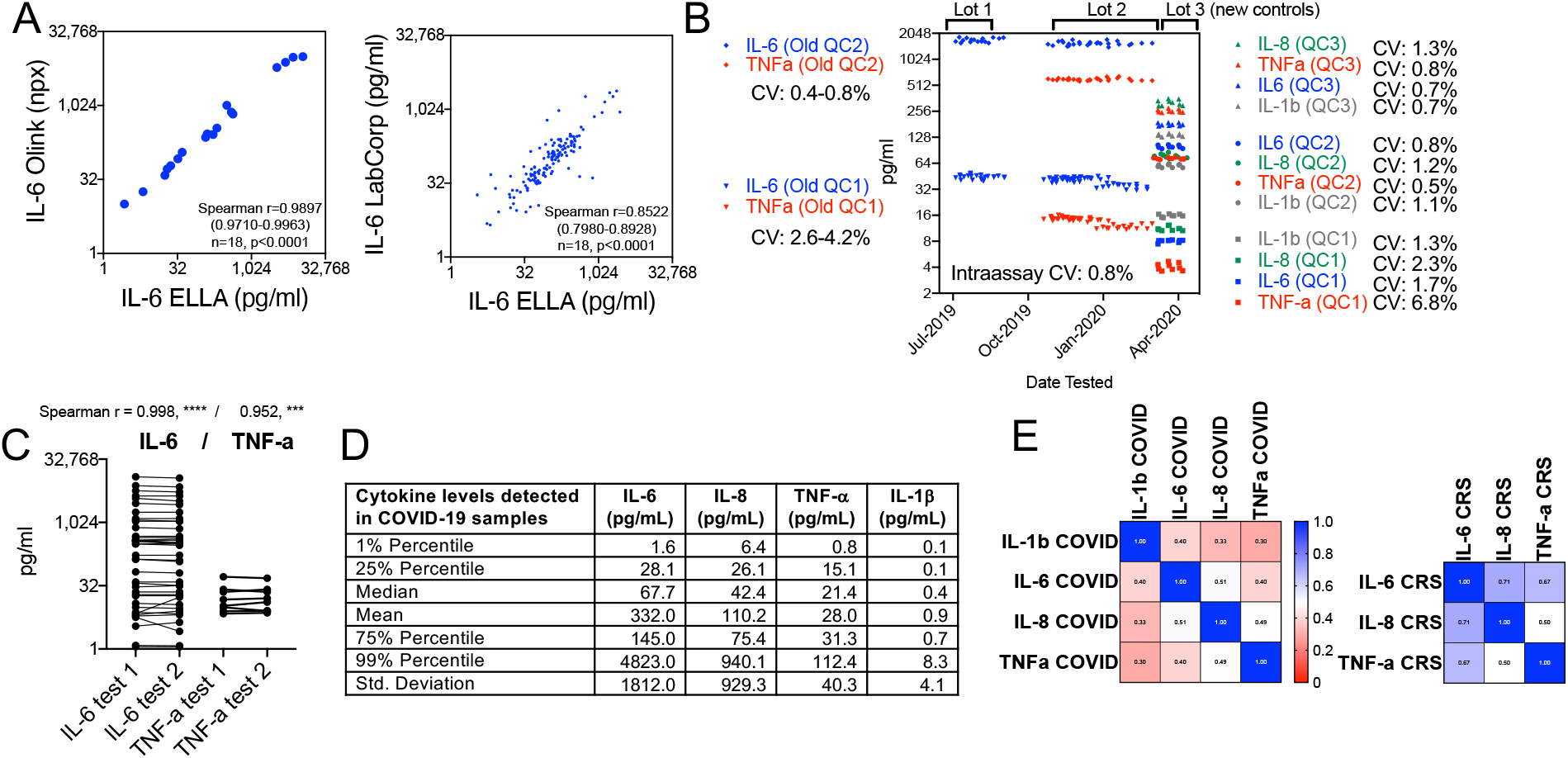
Sensitivity, specificity, and reproducibility testing of the ELLA platform and cytokine levels and correlation observed in COVID-19 specimens. A. Spearman r correlation between IL-6 tested by two other platforms for soluble analyte detection, Olink and LabCorp, using plasma or serum specimens from CAR-T CRS or COVID-19. B. Interassay and intraassay coefficient of variation (CV) of replicates for two recombinant controls used at high or low concentration in each assay for IL-6 and TNF-α using a first set of controls, and for the ELLA panel used in this study using Randox recombinant antigens at three dilution levels. C. Reproducibility testing replicates of the same biological specimen from CAR-T samples, with Spearman r indicated. D. Distribution of each cytokine in all COVID-19 samples tested. E. Correlation matrix of IL-1β, IL-6, IL-8, and TNF-b levels in COVID-19 plasma specimens and in E. multiple myeloma specimens during immunotherapy-related CRS. Scale indicates value of Spearman r correlation. Cytokines levels are less coordinated in COVID-19 than in CAR-T CRS.

**Supplementary Figure S2.**
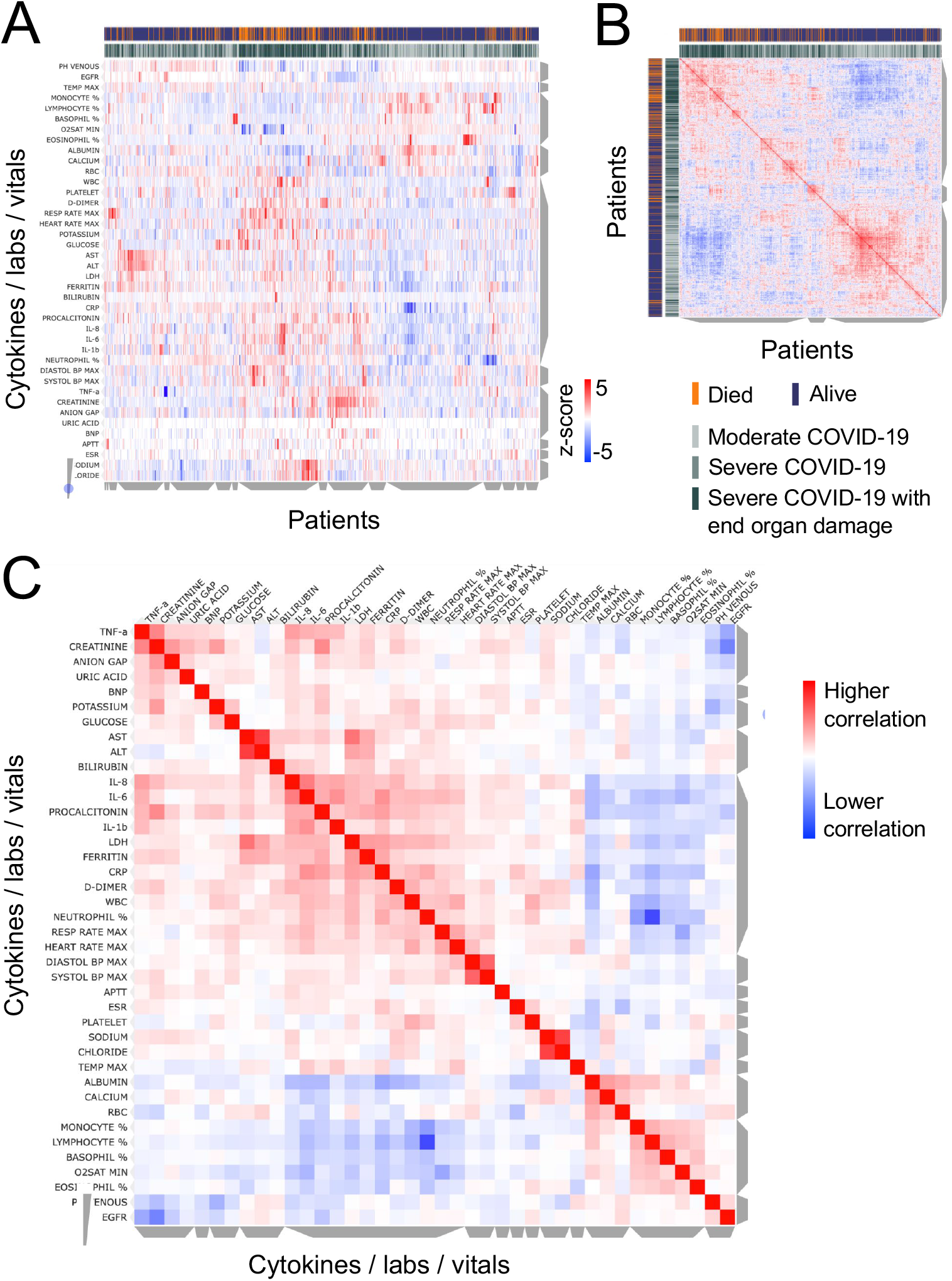
A. Unsupervised clustering of laboratory measurements in a subset of 1069 patients with sufficient available information. On the y axis are vitals and laboratory values after z-scoring, and on the x axis are individual patients, using metrics measured from the time point corresponding to the first ELLA cytokine test. Grey bars on the side of the plot indicate clusters of patients or analytes, where cytokines co-cluster with known severity metrics, such as LDH, CRP, ferritin, D-dimer, but also high neutrophil, platelet and white blood counts. Annotations show patients who died in orange, and maximum severity score achieved in gray shades. B. Similarity matrix of patients based on analytes and measurements, showing two major clusters, with enrichment in patients who died and had more severe COVID-19 on the upper left. C. Similarity matrix of cytokines, lab measurements and vitals, showing IL-6, IL-8, and IL-1β co-clustering with known inflammatory markers such as LDH, CRP, ferritin, and D-dimer, while TNF-α coclusters with organ damage markers.

